# The effect of depth context in the segmentation of the colon in MRI volumes

**DOI:** 10.1101/2020.03.06.20027722

**Authors:** Ezenwoko Benson, Lukas Rier, Isawan Millican, Sue Pritchard, Carolyn Costigan, Michael Pound, Giles Major, Andrew French, Penny Gowland, Tony Pridmore, Caroline Hoad

**Affiliations:** University of Nottingham

## Abstract

Colonic volume content measurements can provide important information about the digestive tract physiology. Development of automated analyses will accelerate the translation of these measurements into clinical practice. In this paper, we test the effect of data dimension on the success of deep learning approaches to segment colons from MRI data. Deep learning network models were developed which used either 2D slices, complete 3D volumes and 2.5D partial volumes. These represent variations in the trade-off between the size and complexity of a network and its training regime, and the limitation of only being able to use a small section of the data at a time: full 3D networks, for example, have more image context available for decision making but require more powerful hardware to implement. For the datasets utilised here, 3D data was found to outperform 2.5D data, which in turn performed better than 2D datasets. The maximum Dice scores achieved by the networks were 0.898, 0.834 and 0.794 respectively. We also considered the effect of ablating varying amounts of data on the ability of the networks to label images correctly. We achieve dice scores of 0.829, 0.827 and 0.389 for 3D single slices ablation, 3D multi-slice ablation and 2.5D middle slice ablation.

In addition, we examined another practical consideration of deep learning, that of how well a network performs on data from another acquisition device. Networks trained on images from a Philips Achieva MRI system yielded Dice scores of up to 0.77 in the 3D case when tested on images captured from a GE Medical Systems HDxt (both 1.5 Tesla) without any retraining. We also considered the effect of single versus multimodal MRI data showing that single modality dice scores can be boosted from 0.825 to 0.898 when adding an extra modality.

## INTRODUCTION

Disturbed motor patterns can lead to various gastrointestinal symptoms (e.g. diarrhoea, constipation and pain) (Wingate, 2003) and imbalances in water secretion or absorption can also affect the nature of the colonic contents and their speed of their transit through the colon. Whilst not life-threatening, persistent symptoms can substantially reduce the quality of life of patients suffering from functional gastrointestinal disorders (Belsey, 2010) and have associated economic costs (Emmanuel and Spinks, 2016). Different techniques have been used to investigate the underlying physiology. Historically, radio-opaque markers and scintigraphy were used clinically to assess meal transit (Szarka and Camilleri, 2012), manometry to study motor patterns (Bampton and Dinning, 2013) and X-ray Computed Tomography (CT) was used to look at anatomical structure. These methods are not ideal, either because they use ionising radiation, or because they disturb normal physiology. They also need to be conducted separately at different times requiring multiple visits and limiting our ability to study the relationship between different parameters.

Recently MRI has emerged as a powerful tool for investigating GI physiology. It allows several key parameters of interest to be measured in a single study and it involves no ionising radiation, allowing multiple imaging examinations, both throughout a day (e.g. to follow a meal) or on different days. Recently MRI assessments of colonic contents, transit, secretions and motility, have provided novel insights into the physiology of constipation (Lam et al, 2016), diarrhoea (Pritchard et al, 2014) and the symptoms associated with irritable bowel syndrome, including pain (Major et al, 2017). Other studies have used MRI measures of colonic volume to study the effects of drugs and laxatives on opioid-induced constipation (Poulsen, 2018), and the effects of different dietary components (Sloan 2018, Bendezu 2017). Importantly the assessment of colonic volume is fundamental to interpreting other physiological measures including motor responses (Lam et al 2016, Pritchard et al, 2014).However, one of the main barriers to the use of MRI for managing functional GI disorders in clinical practice, is the time-consuming nature of the image analysis required, particularly for defining the colonic anatomy and measuring the volume of colonic contents. This problem is particularly complex since the colon extends over may imaging slices within the abdomen, and can vary substantially from person to person and throughout the day, due to bulk motion of contents following a meal or intervention. It would, therefore, be desirable to automate the analysis of colonic volume, reducing the time and expertise needed for manual or semi-automated analyses, reducing subjectivity in the results and making the measurement more clinically applicable.

Segmentation of medical images refers to pixel- or voxel-wise labelling of regions of interest, corresponding to specific organs, tumours, lesions, tissues etc. in 2D or 3D datasets. It is the cornerstone of medical image analysis (Norouzi, 2014), underpinning the development of diagnostic tools, surgical planning and the exploration of anatomy. Manual labelling of large image volumes is time-consuming and challenging due to the problems of visualising and interacting with 3D data, and is prone to uncertainty due to variability between observers (Withey and Koles 2008). Therefore, semi-automatic segmentation is commonly used. For colonic segmentation Sandberg and colleagues (Sandberg et al, 2015) proposed manual delineation of a region of interest around the colon with subsequent unsupervised classification approach to refine the segmentation. While this approach reduced interobserver variability, it took approximately 20 minutes to segment a single dataset, which with the trend towards higher resolution images and temporal datasets will increase the time required for segmentation to unmanageable levels. This burden of data analysis limits the use of these techniques in research and prevents translation to the clinic where image analysis costs must be factored into health economics assessments. In this work we aimed to address these issues by implementing a fully automatic segmentation algorithm using deep machine learning, removing variability due to human observers and bringing the advantage of very short inference times.

Historically, some of the most successful applications of machine learning to pattern detection and classification were implementations of artificial neural networks (ANNs). Originally intended as models of a biological mechanism of perception, ANNs offer a way to approximate solutions to complex problems (Bishop, 2006). ANNs are the successors of the perceptron described by Rosenblatt (Rosenblatt 1958), which was an attempt to model neuronal behaviour, resulting in an output similar to the firing of neurons, given a weighted sum of input ‘stimuli’. The inclusion of a nonlinear activation function allows the perceptron to learn a wide variety of models by adjusting the set of weights applied to the inputs given a set of training data. The combination of these artificial neurons into layered structures resulted in early, shallow ANNs which were successfully used as models for statistical pattern recognition (Bishop, 2006). The rapid development of the field of machine learning and improvements in the performance of computer hardware has enabled the implementation and training of so-called deep Convolutional Neural Networks (CNNs) for use in fully automatic image segmentation. CNNs are a class of supervised and, more recently, semi-supervised machine learning techniques which are well suited to analysing image data. CNNs are composed of 3 main layer types: convolutions, pooling and non-linear. Convolutions are learnable filters centred on a single pixel and applied across the image space. Mathematical operations, usually multiplications, are applied to the surrounding pixels with the results being summed to produce a new value for the centre pixel. Pooling layers collect together inputs from nearby cells or pixels, allowing the network to reduce the spatial or volumetric resolution, saving on memory and computation costs. Max pooling is most commonly used because it favours the pixels with the most intense activation related to the previous convolution. Finally, activation functions are used to introduce non-linearity into the networks; they control the output of a neuron relative to the sum of input activations. Functions used include a variety of piecewise continuous functions such as the sigmoid. In this work rectified linear units (ReLU) (Nair and Hinton, 2010) are used. ReLUs are chosen over continuous functions due to their benefits in training deep networks (Glorot et al, 2011). It is thought that they help prevent the vanishing gradient problem found with the use of sigmoid and other activation functions. ReLU is also a comparatively computationally inexpensive calculation, all negative elements are set to zero and the max operation is applied to the result.

The use of convolutions applied to the output of earlier convolutions allows hierarchical features to be learnt; the first convolutions often learn lower level features such as edges, whereas later convolutions aggregate these to learn more complex, higher-level structures. The training of neural networks involves cycles of repeatedly adjusting the weights and biases in order to approach the desired output. To achieve this, a set of labelled input data is required. The network’s output corresponding to these inputs, given by a forward pass through the network, is evaluated according to a cost function which, in turn, gives a measure of network performance. The result of this cost function can then be used to adjust the network weights during back-propagation, where parameters are adjusted in sequential order beginning with layers closest to the output. Repeating this process will gradually improve the model, leading to a trained network which can then be tested on unseen images. Classically CNNs adopt an architecture of convolutional layers followed by ReLU activations, then pooling layers; this sequence is then repeated multiple times. Each time this occurs the feature maps are downsampled. When applied to the task of classification, the final layer is a fully connected layer, where the number of outputs is equal to the number of classes. These fully-connected layers are so-called as they resemble traditional, completely-connected neural networks, where each node is connected to every node in its neighbouring layers. For semantic segmentation, this fully connected layer is removed and another sequence of layers are appended; here the max pooling operations are replaced with up-sampling layers, which increase the resolution of the features back up to the image scale. Usually, the number of up-sampling layers is equivalent to the number of pooling operations which results in the final output resolution being equivalent to the resolution of the input. The final output is a feature map where each pixel is of a given class.

In the past, the segmentation of medical images has been achieved using a variety of CNN architectures (Ronneberger 2015, Milletari 2016, Zhou 2017). Although successful methods are described in the literature, there is a lack of guidelines on how new datasets should be approached or how machine learning methods could be applied in a clinical or research setting, especially where large datasets are not available and a variety of equipment is used to acquire data. In this work, we demonstrate the automatic semantic segmentation of the ascending colon (AC), transverse colon (TC) and descending colon (DC), using a dataset compiled and segmented for a previous research study (Major et al, 2017). Neither manual nor automatic segmentation of the colon is trivial since colonic motility and varying composition of colon contents cause considerable variability in position, orientation and MRI contrast of the colon, between subjects and over time. Additionally, manual segmentation of individual colon sections which is used as the gold standard, introduces some degree of subjectivity, especially in the exact delineation of the AC-TC and TC-DC boundaries. A human observer tasked to segment the colon will often do so by segmenting individual coronal slices but making use of 3D context and prior knowledge of colon anatomy. Therefore we have compared the segmentation of 2D slices of MRI scans of the abdomen with methods which involve direct segmentation of 3D volumes, as well as a 2.5D approach where only limited 3D context information is available to the segmentation algorithm to explore the effect of depth context on accuracy. We trained the models on data acquired from a single MRI scanner using dual-echo fast field echo sequencing. We then examined the generalisability of the model by using image data from a different source to test our trained models.

## METHODS

Three methods for segmenting colon MRI images are presented below. To ensure a fair comparison between methods all CNNs were trained using a single Nvidia GTX 1080 Ti GPU with 11GB of VRAM. Each network was limited to a maximum of 6 million parameters. All networks were implemented using the PyTorch library version 3 for Python 2.7.

### Dataset

The dataset consists of volumetric MRI scans of the abdomen from healthy volunteers and patients with diarrhoea prominent irritable bowel syndrome (IBS) (Ethical approval 12/EM/0443, National Research Ethics Committee). Each imaging volume was composed of 24 slices in the coronal plane of 256 × 256 pixels. The datasets were acquired using a dual-echo fast field echo sequence resulting in 2 volumes per dataset which were spatially aligned but displayed different contrasts (“echo-1” and “echo-2”; Figure 1). The main dataset used for training and cross-validation was acquired on a 1.5 Tesla Philips Achieva MRI system (Philips dataset) (Major et al, 2017). A dataset acquired using equivalent scanning parameters, but on a 1.5 Tesla GE Medical Systems HDxt MRI scanner (GE dataset), was also used to assess the generalisability of our trained networks to data from different acquisition hardware (Ethical approval 15/LO/0253, National Research Ethics Committee). This GE dataset was never used for training or validation.

**Figure 1:**
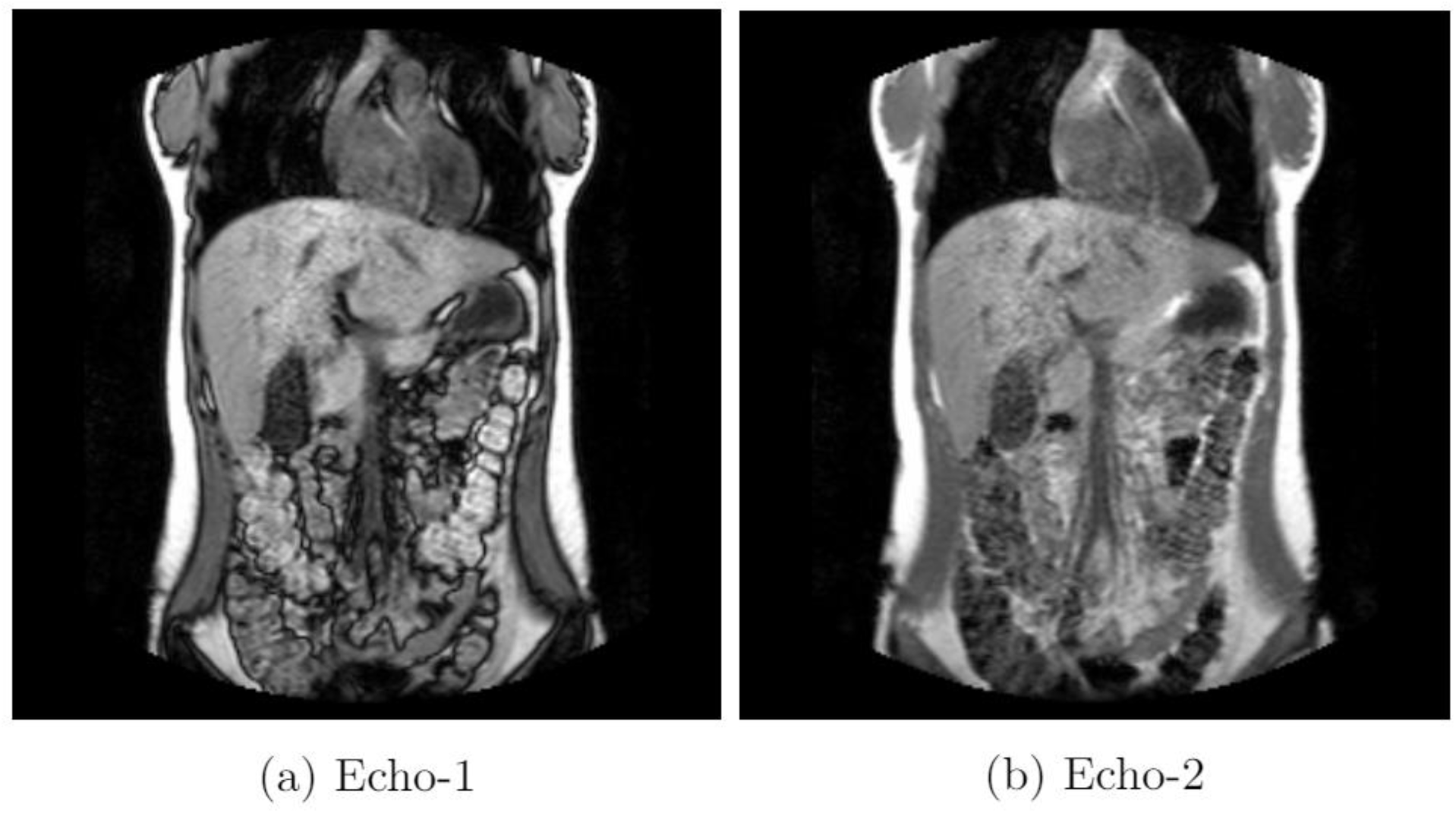
Examples of echo-1 and echo-2 contrasts in a single slice from the Philips Achieva dataset.

The colon regions were manually segmented as described in previously (Pritchard et al, 2014). These regions were then manually segmented into the three foreground (FG) classes: the ascending colon (AC), descending colon (DC) and the transverse colon (TC) using Analyze9 software; the sigmoid colon was not labelled in this dataset. Both echo-1 and echo-2 images were used in combination for the manual and automatic segmentation. The data was from 51 participants, providing up to 7 scans at 2 or 3 visits per subject and consisted of 930 MRI images split into 733 for training, 142 for validation and 55 for testing. Some participants appeared in both validation and test set, but with different scans being used for each set. Thus no scan in the training set was included in the validation or test set.

Figure 2 illustrates the dataset breakdown. No augmentation of the data is used for the training for any of the architectures. We assessed the accuracy of the automatic segmentation of the colon and its sections by computing the Dice overlap coefficient between the model outputs and the manually segmented ground truth. We also calculate IoU and include it for the purposes of comparisons to works which prefer IoU over Dice. The validation set was used to calculate on-the-fly metrics of how models were performing on unseen data.

**Figure 2:**
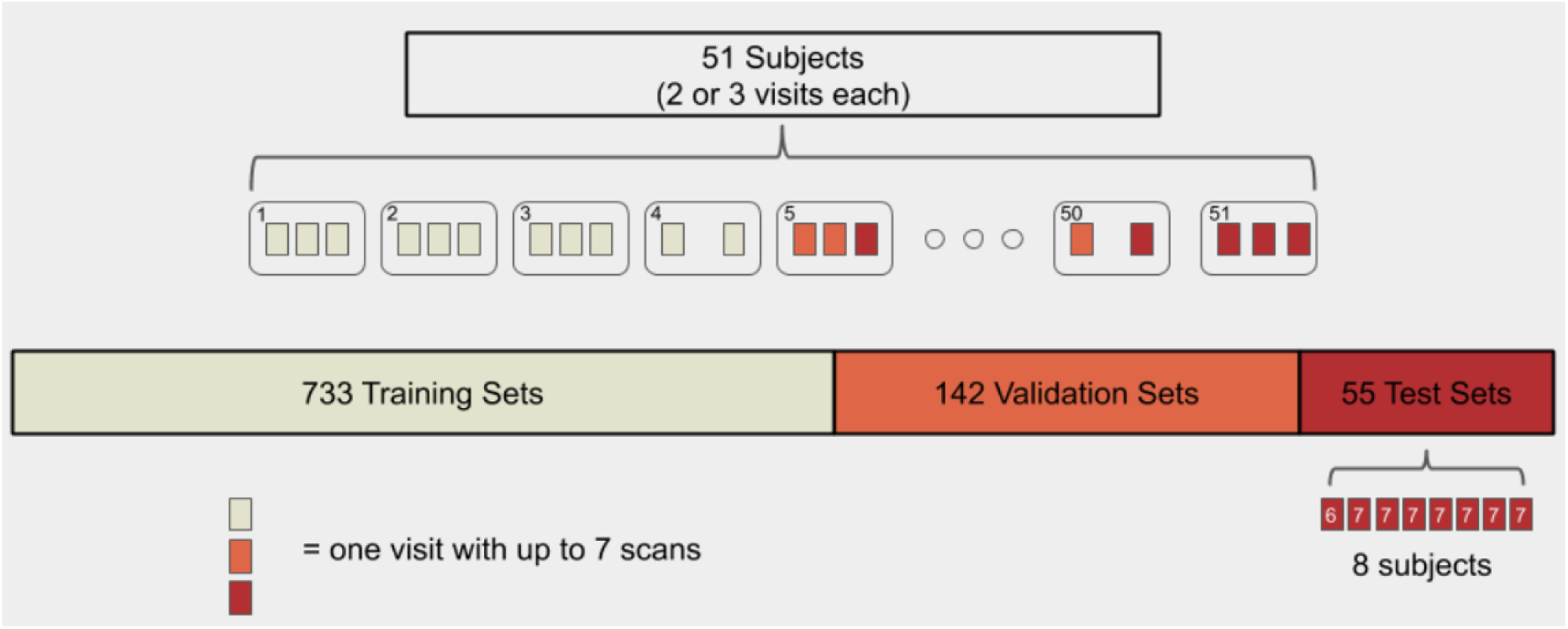
The dataset of 51 participants all of which receives up to 7 scans over 2 or 3 visits. Scans acquired during the same visit are used in the same set; the training, validation or test set. If scans from a participant were used as part of the training set they were not included in the test set.

### Tasks

Models were trained to distinguish between background (BG), AC, TC, and DC. A separate task of binary classification was also explored where the foreground (FG) was a union of separate colon sections. We also calculated a “Whole Colon” (WC average) summary score, which is calculated using the average of AC, TC, DC and BG. Manual segmentation labels were used as the ground truth for both tasks. A comparison of results between the first and second task gave insight into inter-class confusion of the predictions. We also explored the effect of using single contrast MRI volumes compared to multimodal datasets for the tasks of distinguishing between colon regions in each of the 3 architectures.

When comparing the network outputs with the manual segmentations the loss function used is the generalised Dice score (Milletari 2016) for each individual class. The loss was then defined as

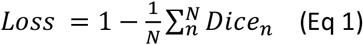

where N is the number of classes and Dice_n_ is the Dice score for the n^th^ class. Dice loss is used as it has been previously shown to train networks which outperform those that use a cross-entropy loss function when solving biomedical image segmentation problems (Milletari 2016). Experimentally we found that training networks with Dice loss results in the same if not better segmentation performance when compared to those trained with cross-entropy loss. Hence we used Dice loss for all our networks. In the next sections we consider the different approaches of the 2D, 2.5D and 3D approaches.

### 2D approach

In this approach, we processed the MRI data using 2D spatial convolutions which were less memory and computationally intensive than their volumetric equivalents. Using spatial convolutions enabled us to have more features in each convolution and allowed for a larger total number of convolutions.

The volumes were presented as a sequence of images (or slices); in order to process the volumes in 2D, we split each volume into images corresponding to its constituent 24 2D slices. Echo-1 and echo-2 were used in combination and were presented to the network as a single entity. In practice, the entity could be seen as an image with two greyscale channels.

#### Model architecture

The same 2D network architecture was used for each echo experiment with differing inputs. The architecture used was a stacked hourglass structure (Newell et al, 2016). The hourglass followed a classic semantic segmentation architecture: it was separated into two sections, encoder and decoder as seen in Figure 3. The encoder segment contains bottleneck blocks (He et al, 2016) followed by max-pooling to perform spatial downsampling. At each spatial resolution, one bottleneck block was used until the lowest resolution. The network contains featuremaps at four different spatial resolutions which is achieved through four max pooling operations, which reduced the spatial resolution of the feature maps to 16 × 16.

**Figure 3:**
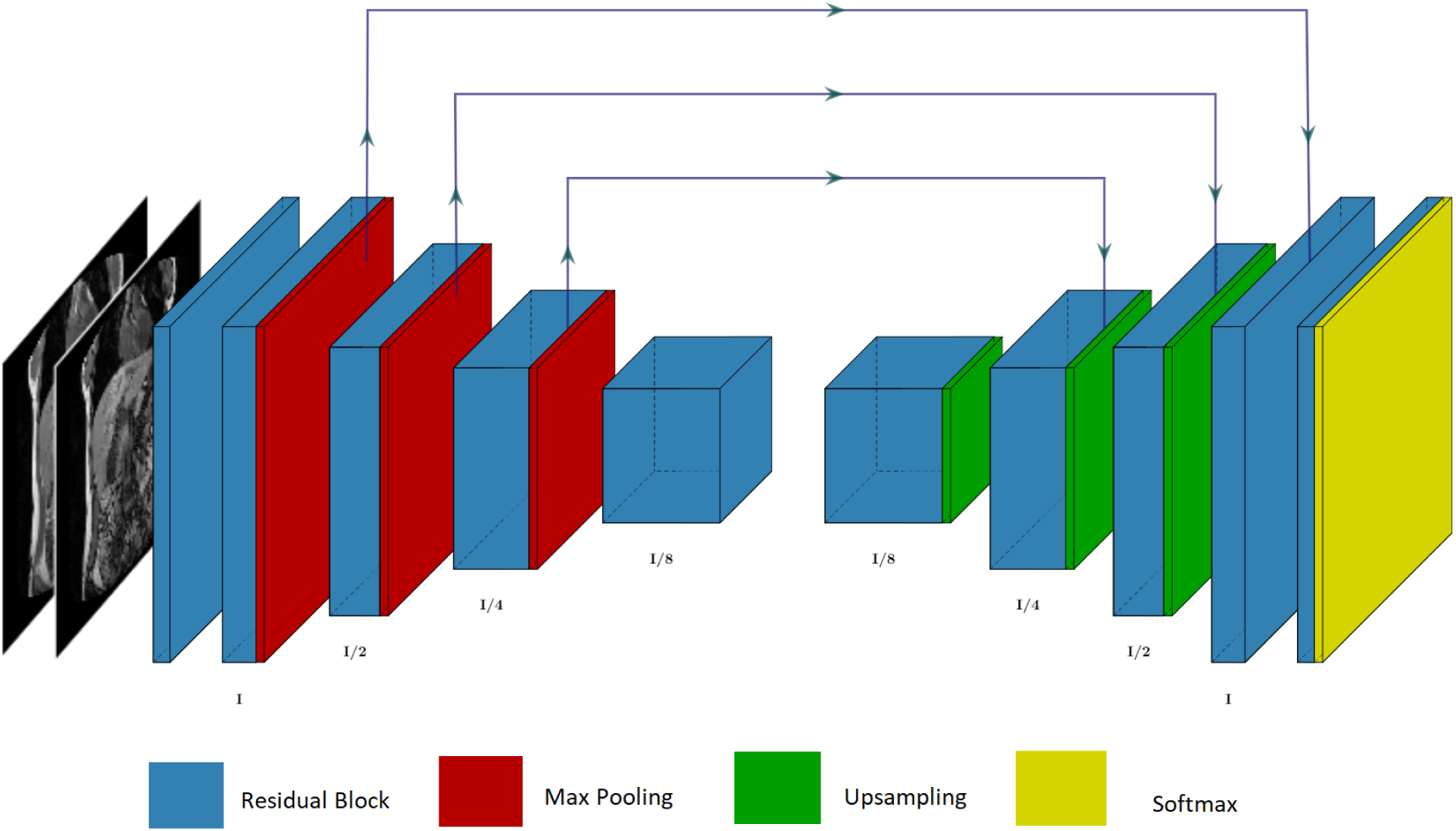
The 2D hourglass architecture used in this paper.

The decoder segment of the network also contained bottleneck blocks but, conversely, each was followed by nearest-neighbour upsampling; four were used in total to restore the output to the original input image’s resolution. The hourglass used skip connections between bottleneck blocks with equivalent resolutions in the encoder and decoder. The skip connections were concatenations of the feature maps followed by a 1×1 convolution to reduce the number of feature maps by a half. This stops the decoder from doubling in size in comparison to the encoder.

#### Training

As mentioned above the MRI datasets were split into slices, and the network was trained on a large set of slices. A mini-batch is a very small subset of the training data which is presented to the network. Presenting the network with more than a single instance of the training data allows the network to learn features across the training data more reliably. In separate training instances, mini-batch sizes were varied and a mini-batch size of six was found to be the most optimal. ADAM (Kingma and Ba 2014), a stochastic gradient descent algorithm variant, was used to train the network at a learning rate of 10^−5^, which was selected empirically. The test set was used to compute the final results shown in the quantitative analysis section.

### 2.5D approach

This approach used the same methodology outlined in the 2D approach: volumes were separated into slices, but each slice was given as input to the network as a triplet consisting of the preceding slice, current slice and proceeding slice.

Anatomically, these are the anterior, current and posterior MRI slices. For the first slice of a volume, the preceding slice was left as empty data, with its corresponding ground-truth label set as BG. The last image in a volume had its proceeding slice set in the same way.

As with the 2D approach, experiments were carried out with echo-1, echo-2 and both combined, therefore the input used for the network was a six-channel image volume of size 6 × 256 × 256, which contains two corresponding triplets.

#### Model Architecture

The architecture of the network differed only in its input layer. In the 2.5D network, neighbouring slices were input to the network as separate channels so each input slice was no longer a single greyscale image, but rather was a three greyscale triplet image. In the case that both echo-1 and echo-2 were used as input, the input would contain 6 (3 × 2) channels as illustrated in Figure 4.

**Figure 4:**
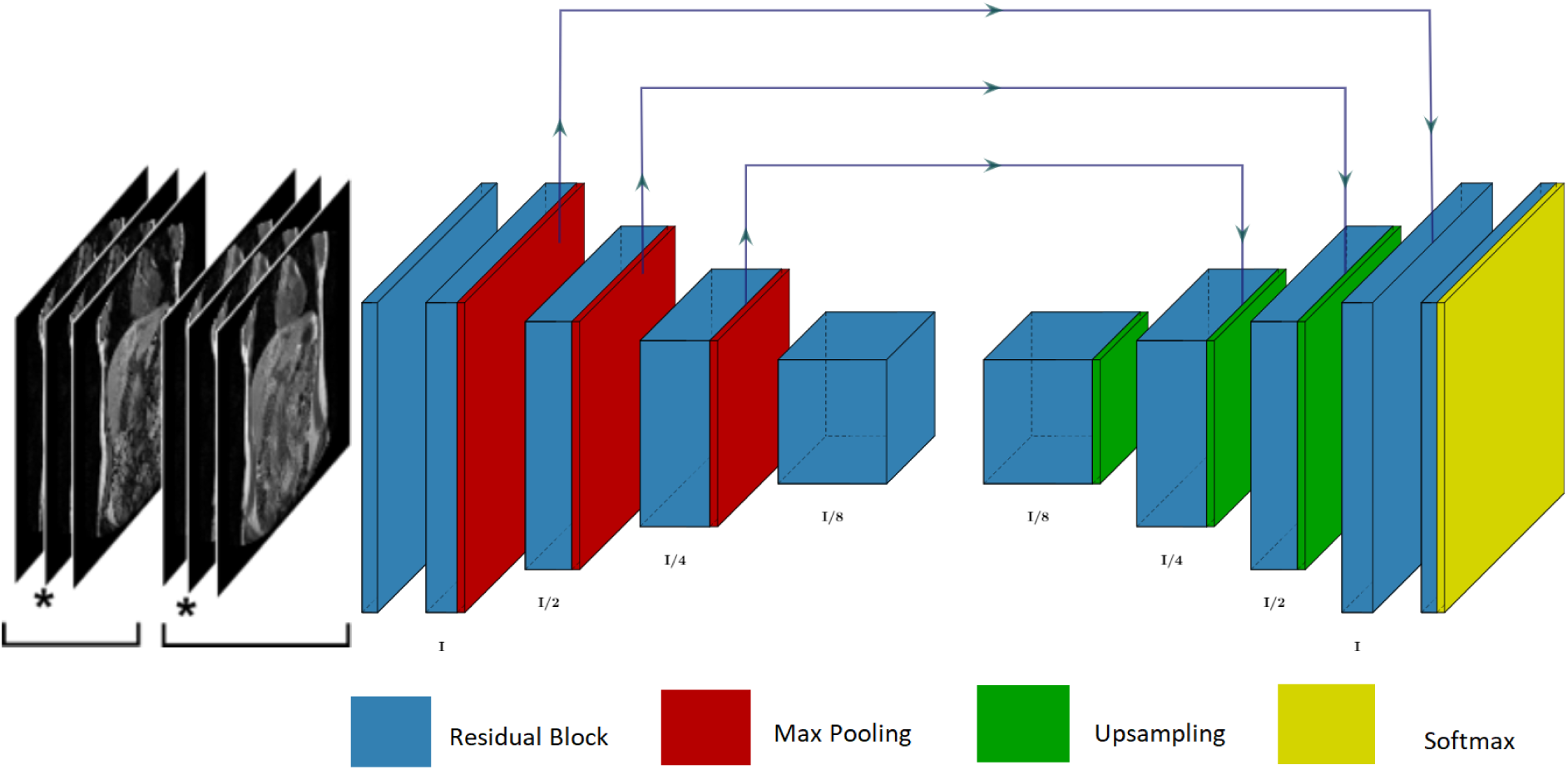
The 2.5D hourglass architecture used in this paper. The asterisk on the displayed input image indicates which slice of the input is segmented. The 2.5D network only segments the middle slice but uses surrounding slices for context

#### Training

The training method used for the 2.5D network was identical to the method presented in the 2D approach training subsection.

### 3D approach

For this approach, we explain the use of volumetric convolutions in an encoder-decoder architecture to achieve semantic segmentation of the individual sections (AC, TC, DC). In 3D we compared the segmentation performance of the trained models when using image volumes consisting of a single contrast channel – either echo-1 or echo-2 - or combined images containing both contrast channels for training. The 3D convolutions provided more contextual information regarding the structure of the colon, but had higher memory requirements, thus reducing the possible depth of the network for computational reasons. In addition, the use of 3-D kernels greatly increased the number of network parameters.

#### Model Architecture

Our network architecture was the same as the 2D hourglass except that all spatial convolutions were replaced with volumetric convolutions. The network was modified to accept two echo contrasts as inputs: each echo image was treated as a channel of a two-channel image. The model consisted of an encoder/decoder structure, where features were extracted using trainable convolutional kernels.

Volumetric max-pooling with stride 2 was used to reduce dimensionality within the encoder structure, while nearest neighbour upsampling operations were used to match the output resolution with the input images. Small scale features were extracted from the shallow parts of the network, while larger-scale features were extracted from deeper layers. In order to preserve small scale features from the encoding layers, skip connections between encoding and decoding layers were implemented. Unlike in the 2D and 2.5D architectures we did not use concatenations followed by 1 × 1 convolutions for our skip connections. Instead, we used element-wise summation to save on VRAM (GPU memory) and network parameters. Residual connections between feature layers prevented the vanishing of gradients during back-propagation (He 2016). We used ReLU as the activation function, and a softmax layer was used to normalise the output. An overview of our final architecture is given in Figure 5.

**Figure 5:**
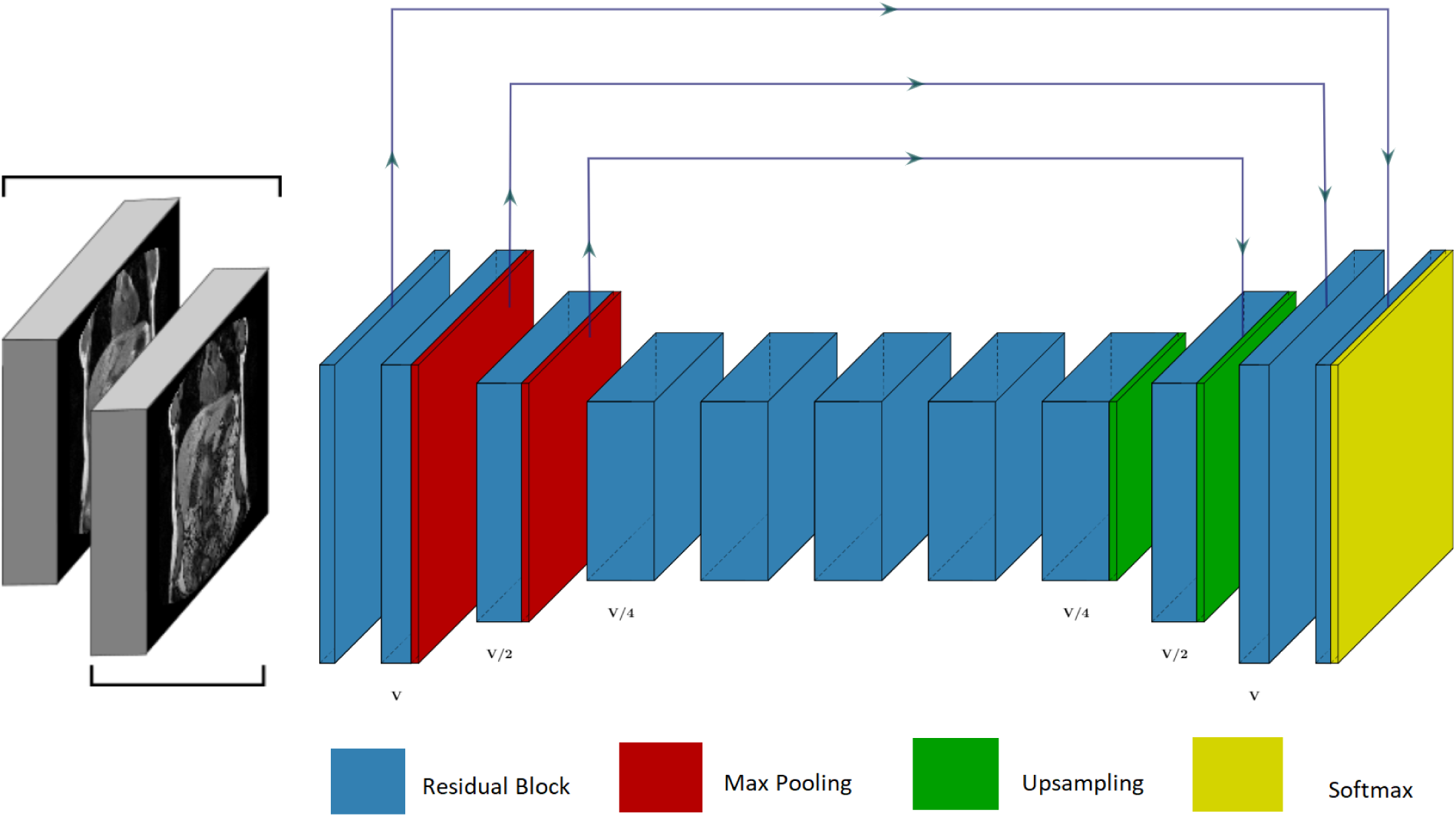
The 3D hourglass architecture used in this paper.

#### Training

We trained 3D models on a single Nvidia GTX 1080 Ti graphics card, where each model used approximately 11GB of VRAM. A mini-batch size of 1 was used; larger mini-batch sizes were not investigated due to memory constraints. The training of the model-parameters was achieved using the ADAM optimiser with a learning rate of 10^−6^.

## RESULTS

### Single vs Dual Contrast

Given the two available contrasts, we trained separate models using either echo-1 or echo-2 images as input using WC ground truths. Figure 6 shows the Dice score during the training of the single contrast modalities and the combined contrast modality. When inputting a single modality, echo-2 obtained significantly higher accuracy than echo-1, whilst dual-modality contrast input provided the best accuracy and trained the fastest. As can be seen in Figure 7, the model using echo-2 contrast model was more successful at segmenting the colon than the echo-1 model. However the highest Dice score was achieved when the image volumes with different contrasts were combined into a 4D matrix, with the fourth dimension representing the contrast channels. Given this result, all subsequent models were trained using dual-contrast image volumes as training data.

**Figure 6:**
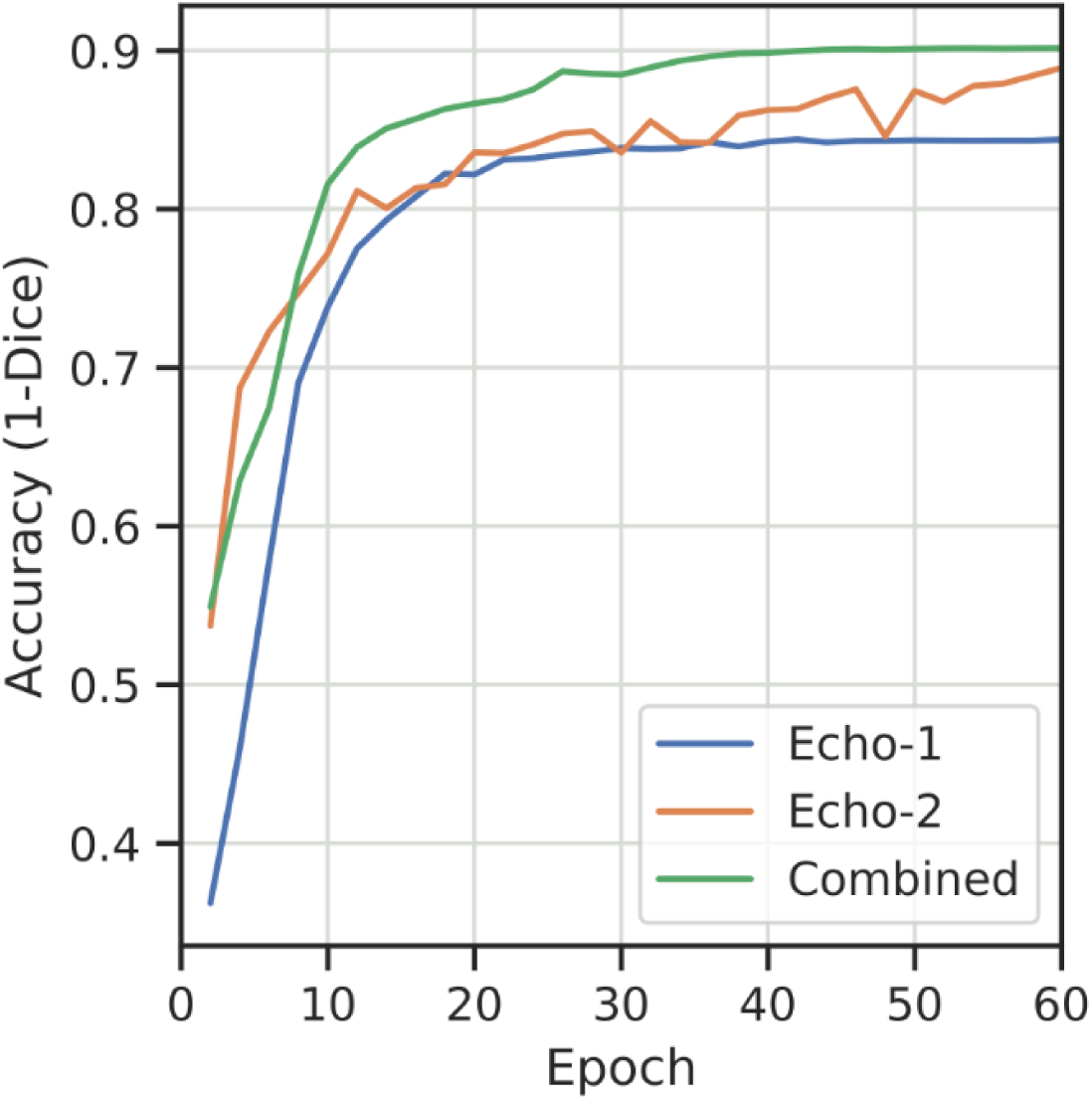
Plot showing the change in segmentation accuracy during training time for input volumes with a single contrast (Echo-1 or Echo-2) and for dual contrast inputs.

**Figure 7:**
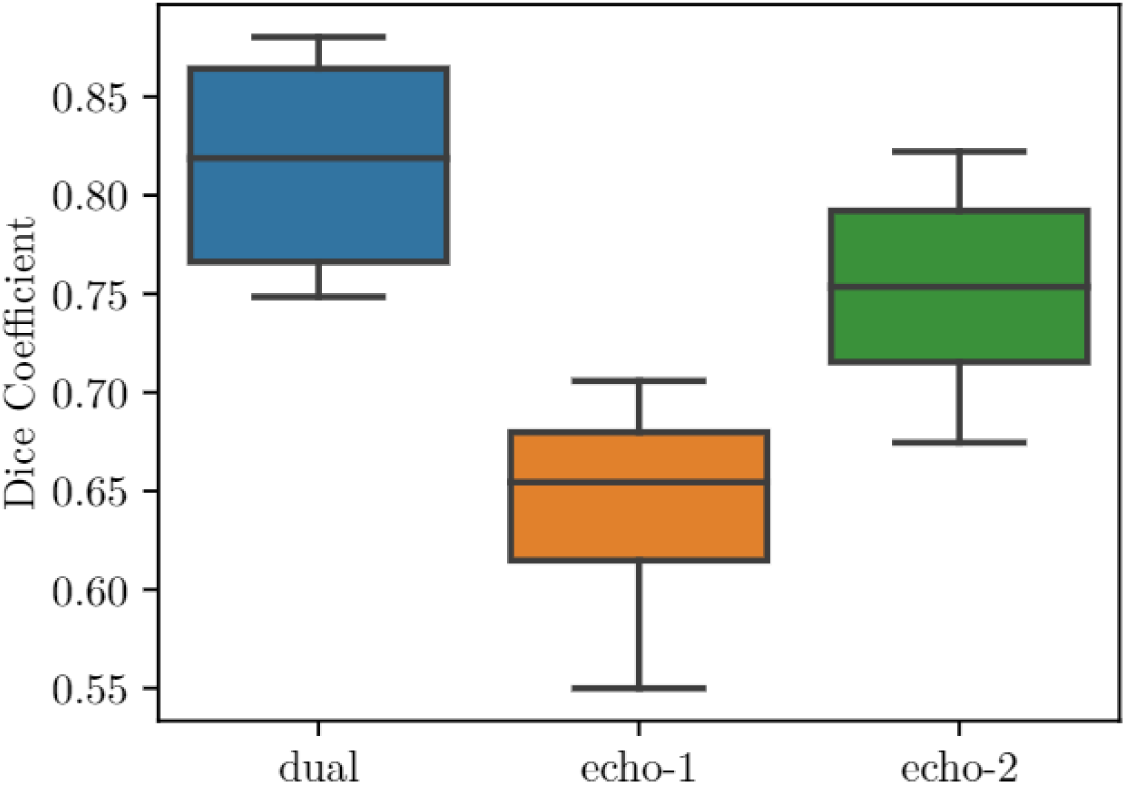
Box plots showing the 10^th^, 25^th^, 50^th^, 75^th^, and 90^th^ percentile of the Dice scores achieved by 3D models trained on dual-contrast images, echo-1, and echo-2 images respectively. Results obtained on the Phillips Achieva test data.

In the following tables we break down the tasks based on the subject of the segmentation. WC refers to the segmentation of individual colon sections and is calculated as the average accuracy of the 4 classes BG, AC, DC and TC. A measure of accuracy is also provided for each section of the colon. The final row, “Binary”, presents the accuracy of the network when trained to perform binary segmentation into two classes, simply Colon or BG.

## Qualitative Analysis

Quantitative metrics provide a useful method for comparing competing architectures, but give few insights into the semantic strengths and weaknesses of a network. In the following sections, we show visual results from each network and discuss some common issues that arise from the network predictions. In each Figure, the contour indicates the network prediction, whilst the solid overlay represents the ground truth. Only a single channel of the dual echo input is shown. For each input image, we show the corresponding network prediction for binary as well as individual colon sections; the binary and individual colon segmentations are separated in the figures given.

### Phillips Achieva

Figure 8 displays examples from a well-segmented case, the average Dice score achieved across the three networks for this image was 0.90. In such cases, the networks were able to produce segmentations with only a small amount of class confusion and FG-BG confusion. The contours were relatively smooth and most colonic regions were captured.

**Figure 8:**
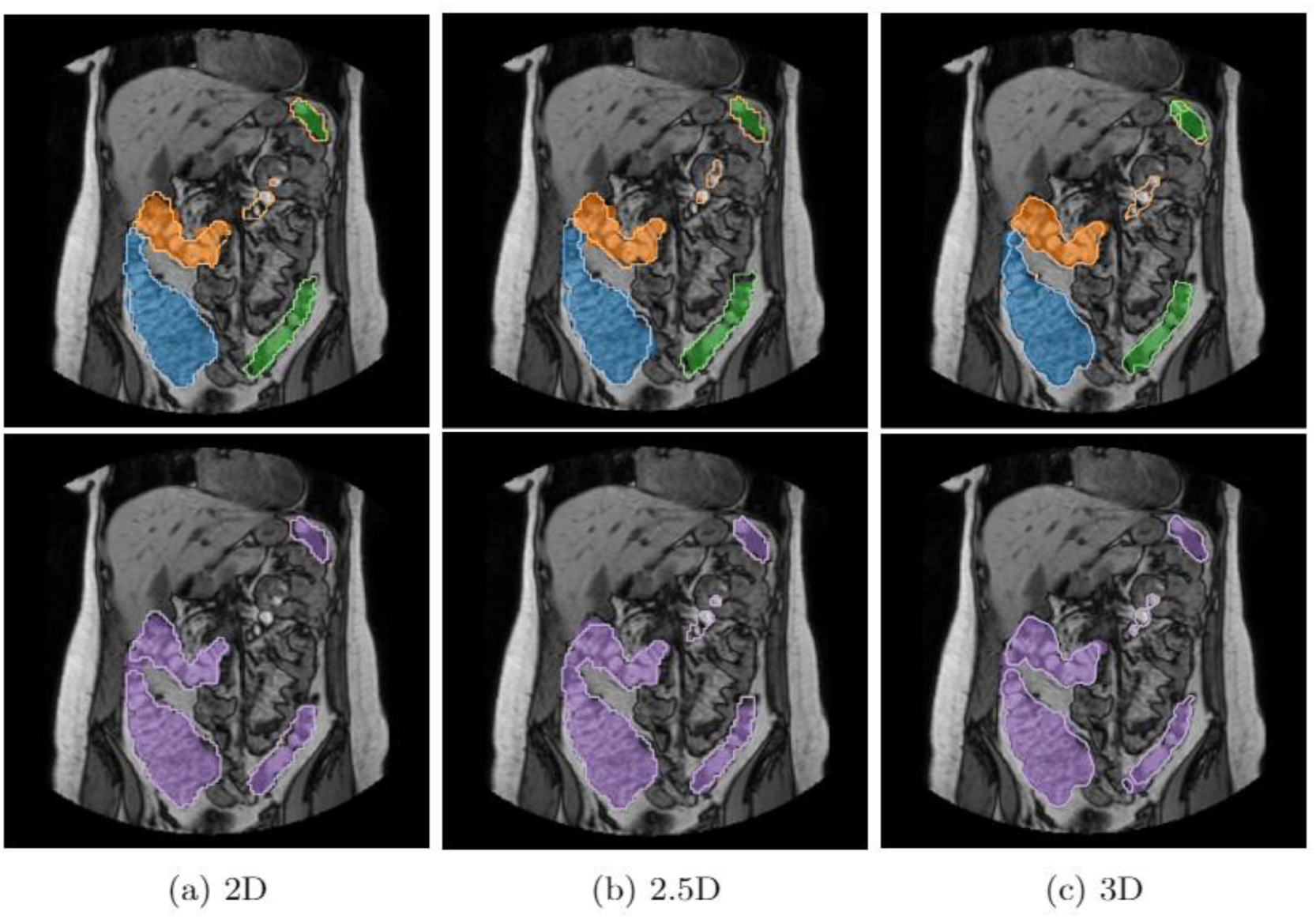
Examples of network predictions on a volume with high accuracy. Coloured regions denote the ground truth with the blue, orange and green corresponding to AC, TC and DC respectively. Slices with purple labels are the corresponding binary segmentations. The network outputs are delineated by the thin contour lines.

In Figure 9 we present network outputs on a volume where all networks perform only averagely well. The average Dice score for this volume was 0.836. In these cases large contoured regions of FG-BG misclassification were common. Class confusion between colon segments was also common. Well defined regions with high contrast which fall between the AC and DC were often misclassified as TC. Finally, in Figure 10 we present network predictions with low accuracy. The contours of each network were much less smooth with significant regions of the colon being missed altogether. Contours often overlapped with BG pixels or exist as a sub-region of the ground truth. In these more difficult cases, the networks had differing disadvantages. The 3D network often made predictions around regions which did not show the contrast expected in the colon, as seen in Figure 10(c). Upon further investigation these regions often contained colon further along the volume or in preceding slices. Therefore, we hypothesise that the 3D network attempted to segment colon regions it expected to appear in certain positions. In the more difficult cases, the 2D and 2.5D network suffered similar disadvantages. Both networks missed many colon regions which present in each slice whilst the 2.5D network seemed to be more prone to misclassification upon inspection of the images.

**Figure 9:**
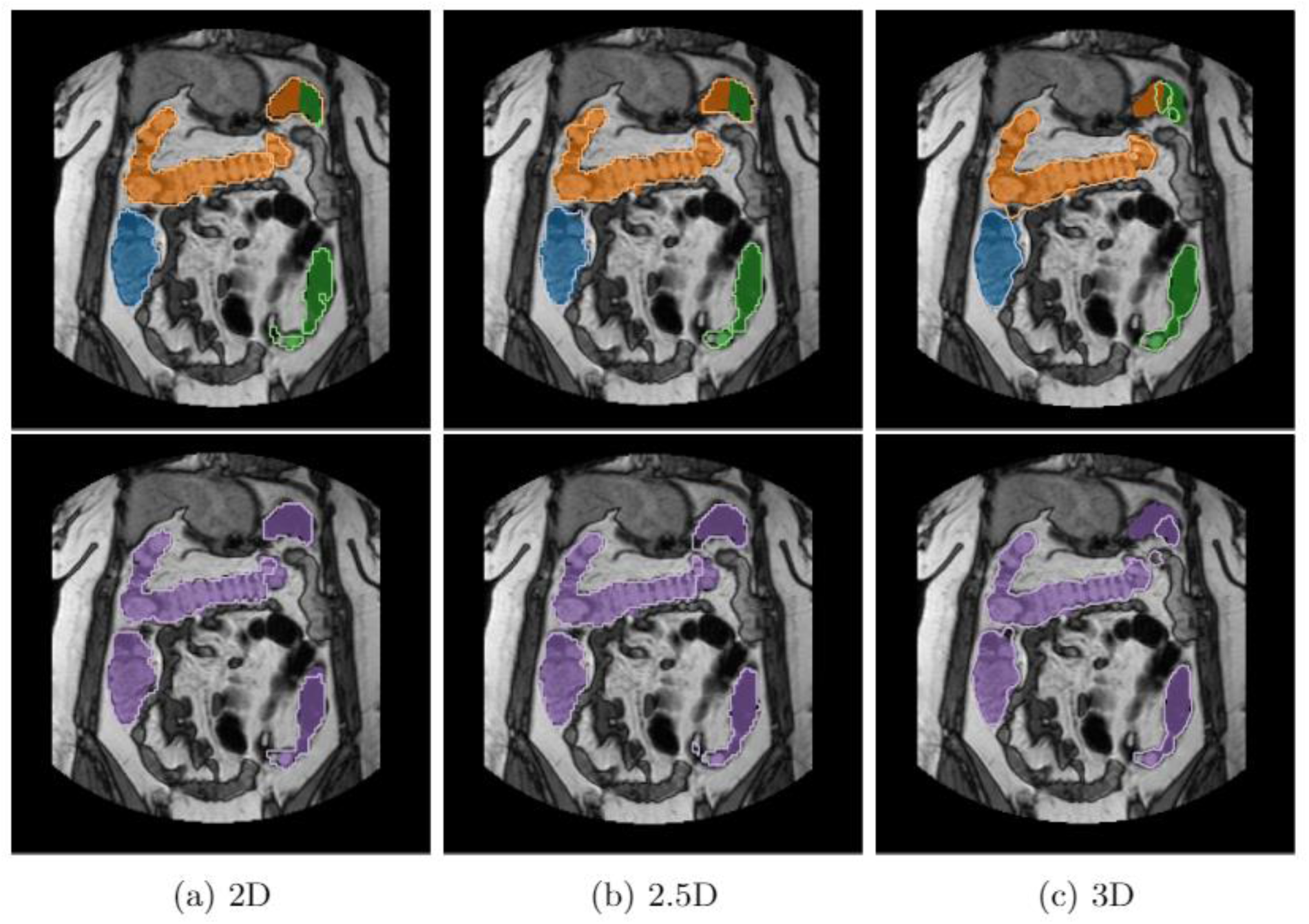
Examples of network predictions on a volume with only average accuracy. Coloured regions denote the ground truth with the blue, orange and green corresponding to AC, TC and DC respectively. Slices with purple labels are the corresponding binary segmentations. The network outputs are delineated by the thin contour lines.

**Figure 10:**
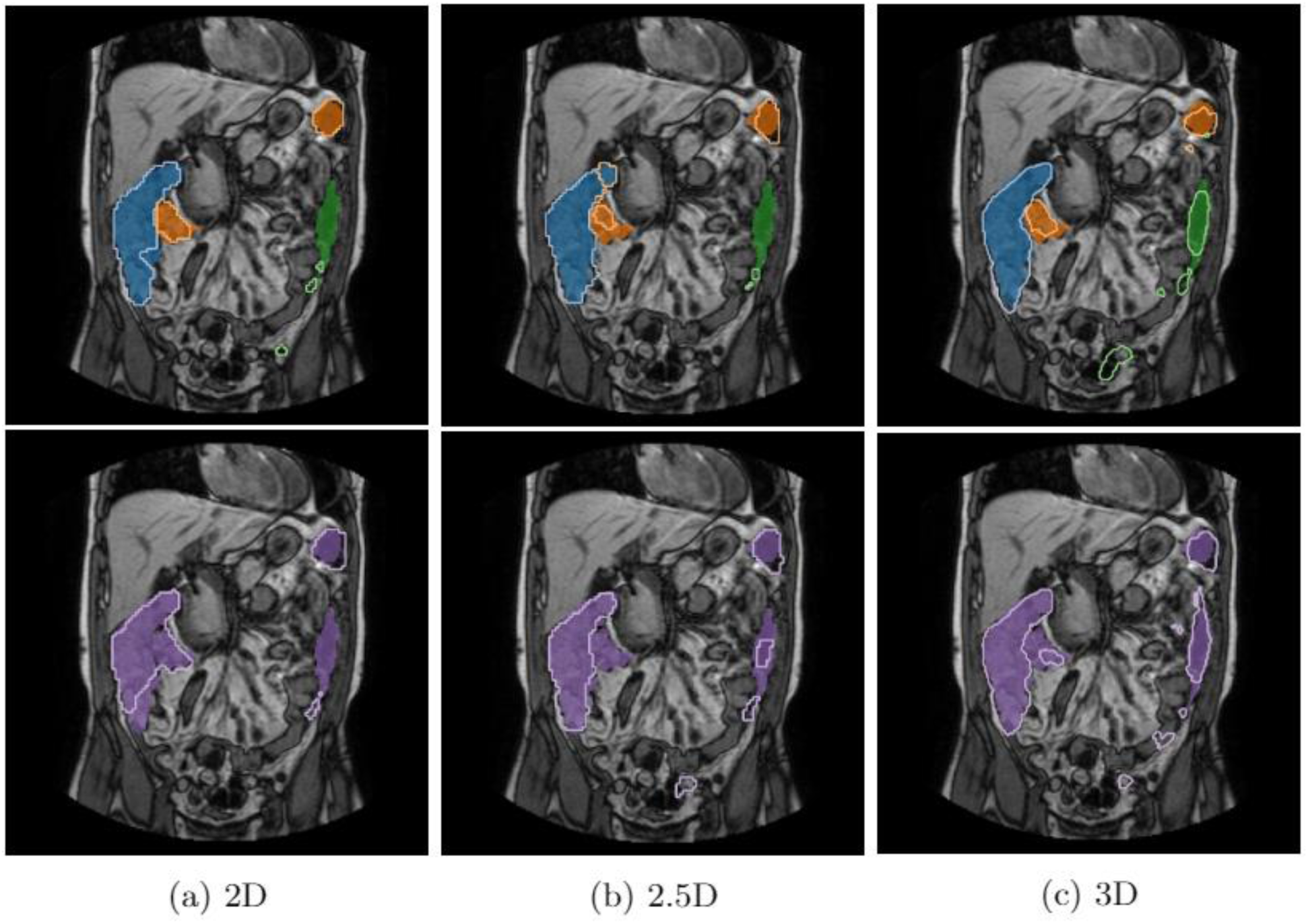
Examples of network predictions on a volume with a low accuracy. Coloured regions denote the ground truth with the blue, orange and green corresponding to AC, TC and DC respectively. Slices with purple labels are the corresponding binary segmentations. The network outputs are delineated by the thin contour lines.

The colon region which was most often misclassified as the BG was the descending colon. Below the descending colon exists the sigmoid colon which has very similar contrast information to the descending colon, but the sigmoid colon was not defined in the ground truth manual segmentations, and the endpoints of each section are ill-defined. Since the sigmoid colon was treated as BG we hypothesise that the networks struggled to differentiate between the two regions causing a BG misclassification.

### GE Dataset

Since this dataset did not form part of the training set, and transfer learning was not used, we expected results here to be worse than those above. Overall results look acceptable, but class confusion between the colon regions is high and most notably FG-BG class confusion is common, as shown in row 2 of Figure 11.

**Figure 11:**
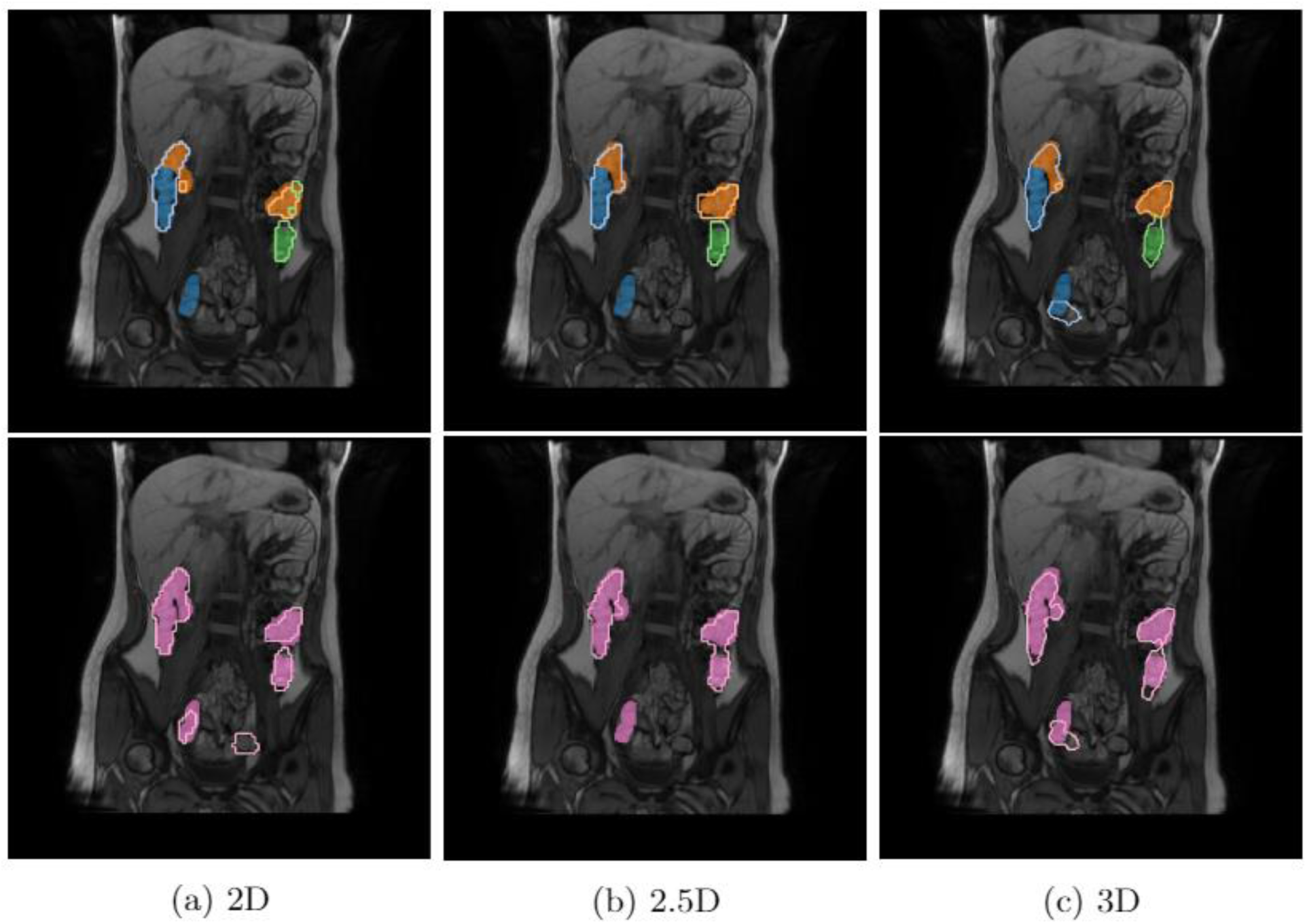
Examples of network predictions on a volume with the highest accuracy between networks on the GE scanner.Coloured regions denote the ground truth with the blue, orange and green corresponding to AC, TC and DC respectively. Slices with pink labels are the corresponding binary segmentations. The network outputs are delineated by the thin contour lines.

Misclassified voxels of FG and BG occurred in all segmentation scenarios; however, we found that regions which clearly had a different appearance to that found in the Phillips dataset contained significantly more misclassified voxels (Figure 12). We hypothesise that this occurred due to the difference in contrast information between scanners. We found artefacts not present in the training set in many of the scans in the GE scanner dataset, and often these artefacts were misclassified as regions of colon. Since the Philips scanner dataset did not contain many of these artefacts, the network was not trained to deal with such noise. This is important since different scanning methodologies are likely to generate their own unique artefacts, but with transfer learning we may be able to reduce the effect of these.

**Figure 12:**
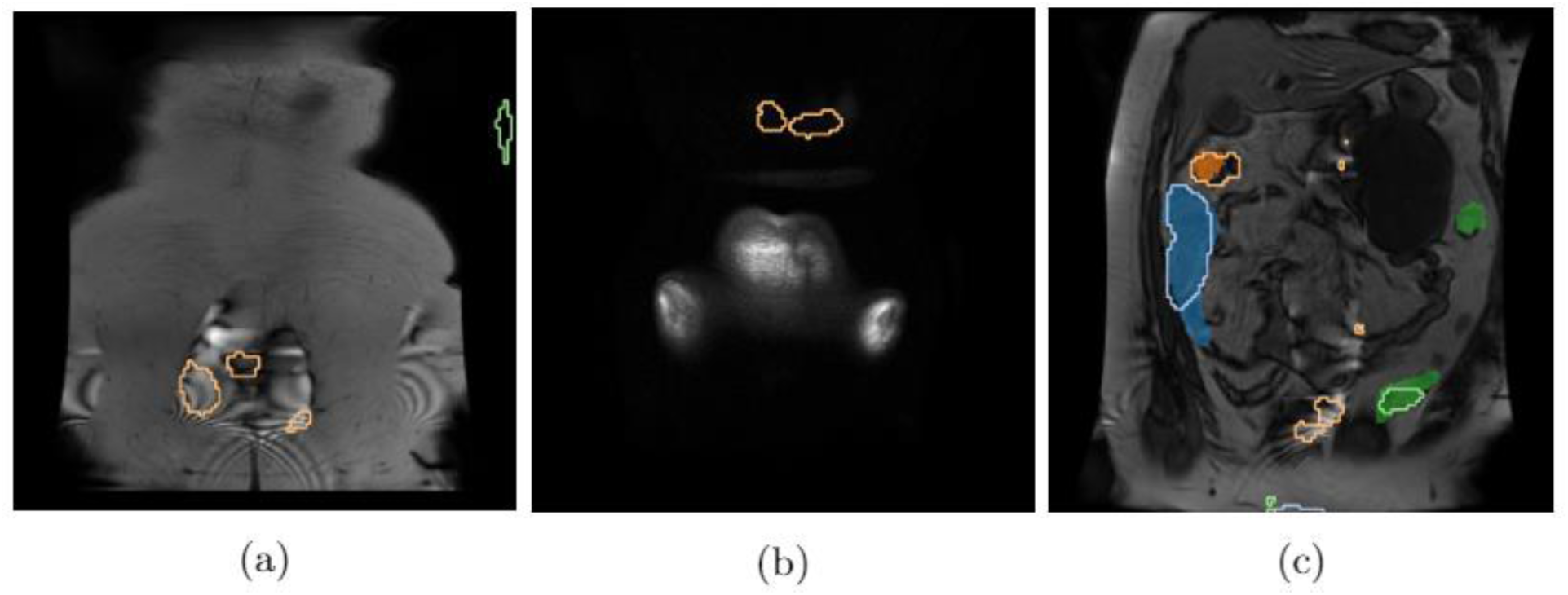
Noisey 2D network predictions in coronal slices. (a) and (b) do not contain any colon regions. All images show incorrect predictions whilst (a) and (c) show artifacts within the MRI which cause misclassification. Coloured regions denote the ground truth with the blue, orange and green corresponding to AC, TC and DC respectively. The network outputs are delineated by the thin contour lines.

## Quantitative Analysis

### 2D approach

The 2D approach was trained for 150 epochs. Training lasted approximately 16 hours and required around 5.3GB of VRAM. Table 1 shows the results for the 2D network for each of the sections of the colon, as well as the whole colon segmentation.

**Table 1:** Segmentation accuracy results obtained for the 2D network applied to test subsets of the Philips and GE datasets. WC denotes the average segmentation accuracy over all four individually segmented classes (including BG). AC denotes segmentation of the ascending colon, DC the descending colon, and TC the transverse colon. ‘Binary’ segmentation refers to the task of segmenting the regions containing the colon from the BG.

| Class | IoU | Dice |
| --- | --- | --- |
| **Philips Achieva** |  |  |
| WC Average | 0.754 | 0.794 |
| AC | 0.736 | 0.785 |
| DC | 0.606 | 0.669 |
| TC | 0.683 | 0.725 |
| Binary | 0.849 | 0.887 |
| **GE Medical Systems HDxt** |  |  |
| WC Average | 0.461 | 0.525 |
| AC | 0.386 | 0.443 |
| DC | 0.257 | 0.325 |
| TC | 0.216 | 0.258 |
| Binary | 0.700 | 0.747 |

### 2.5D approach

The 2.5D approach was also trained for 150 epochs. Training time and VRAM usage were identical to the 2D approach. However, whilst the input was temporarily held in memory, the RAM usage was slightly higher than in the 2D approach. This is due to the difference in channel sizes of the inputs; the 2.5D network has three times as many input channels which must be held in memory. This only makes the input channel size of the first layer in the network three times larger. Table 2 shows the results for the 2.5D network on the test set.

**Table 2:**
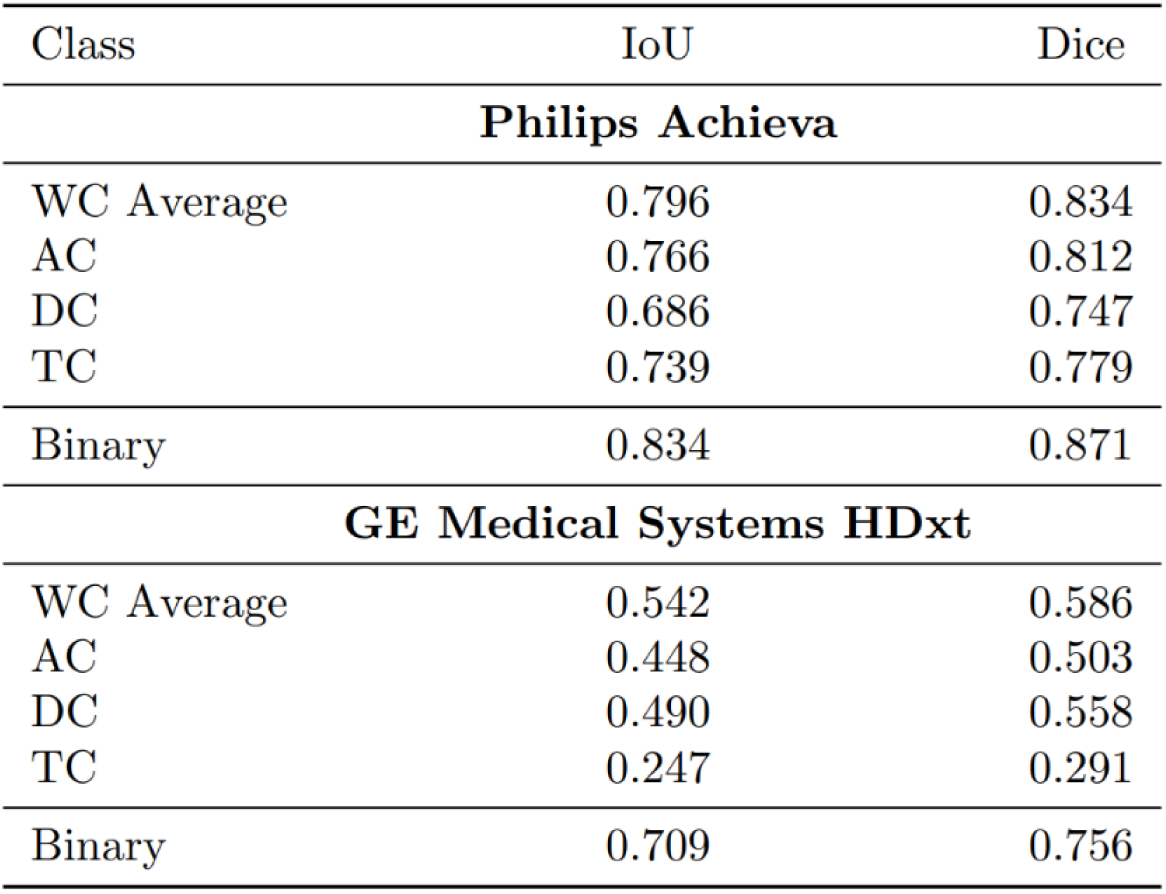
Segmentation accuracy results obtained for the 2.5D network applied to test subsets of Philips and GE datasets. WC denotes the average segmentation accuracy over all four individually segmented classes (including BG). AC denotes segmentation of the ascending colon, DC the descending colon, and TC the transverse colon. ‘Binary’ segmentation refers to the task of segmenting the regions containing the colon from the BG.

| Class | IoU | Dice |
| --- | --- | --- |
| **Philips Achieva** |  |  |
| WC Average | 0.796 | 0.834 |
| AC | 0.766 | 0.812 |
| DC | 0.686 | 0.747 |
| TC | 0.739 | 0.779 |
| Binary | 0.834 | 0.871 |
| **GE Medical Systems HDxt** |  |  |
| WC Average | 0.542 | 0.586 |
| AC | 0.448 | 0.503 |
| DC | 0.490 | 0.558 |
| TC | 0.247 | 0.291 |
| Binary | 0.709 | 0.756 |

### 3D approach

For the approach using the 3D architecture, we trained our models for 150 epochs. The VRAM usage at training time was 11GB. Results for this network can be seen in Table 3.

**Table 3:** Segmentation accuracy results obtained for the 3D network applied to test subsets from Philips and GE datasets. WC Average denotes the average segmentation accuracy over all four individually segmented classes (including BG). AC denotes segmentation of the ascending colon, DC the descending colon, and TC the transverse colon. ‘Binary’ segmentation refers to the task of segmenting the regions containing the colon from the BG.

| Class | IoU | Dice |
| --- | --- | --- |
| **Philips Achieva** |  |  |
| WC Average | 0.863 | 0.898 |
| AC | 0.848 | 0.890 |
| DC | 0.766 | 0.823 |
| TC | 0.842 | 0.882 |
| Binary | 0.900 | 0.933 |
| **GE Medical Systems HDxt** |  |  |
| WC Average | 0.728 | 0.770 |
| AC | 0.732 | 0.785 |
| DC | 0.606 | 0.674 |
| TC | 0.585 | 0.627 |
| Binary | 0.837 | 0.882 |

### Quantitative Summary

Summarizing the results in Tables 1-3: the 3D network is the best performing network with an overall dice score of 0.898 and a dice score of 0.933 for whole colon binary classification. When looking at WC average (overall segmentation) the 2D network was outperformed by the 2.5D network by 0.040 and the 3D network outperforms the 2.5D by 0.064. For binary classification of FG and BG the 3D network performs the best followed by 2D and 2.5D which perform very similarly. The 3D network, which performed best overall also performs best at segmenting the AC, TC and DC sections individually. These statements are true for both IoU and Dice.

## Generalisability to other datasets

The results for the GE data set are shown in Tables 1, 2 and 3, beside the results from the original Philips dataset for comparison. In general, running models on the GE data led to lower segmentations accuracies. However, it is worth noting the model trained to segment the colon as a whole (Binary) performed significantly better than the models trained to segment individual colon sections. Each network’s relative ability to segment sections of the colon remained the same between scanners: 2D, 2.5D and 3D networks achieved Dice scores of 0.525, 0.586 and 0.770 for WC Average segmentation respectively. The difference in contrast information appears to affect the 2D and 2.5D networks the most. BG-FG class confusion is very high amongst the networks which probably accounts for the large decrease in performance (see Figure 13). However the confusion between BG and TC is very similar to the DC which was not seen with the original Philips scanner data (see Figure 14). Example output images in the qualitative analysis section show noisy prediction where BG pixels were predicted as colonic regions - this could be due to artefacts and change in contrast information between the scanners. Figure 12 (a) shows an example of a ringing artefact being wrongly classified as being part of the transverse colon. Given that a similar texture of parallel dark and light stripes also delineates the folding tissue between the colon’s haustra (pouch-like structures forming the colon), one might speculate that this texture was learned by the network as a useful feature for colon segmentation.

**Figure 13:**
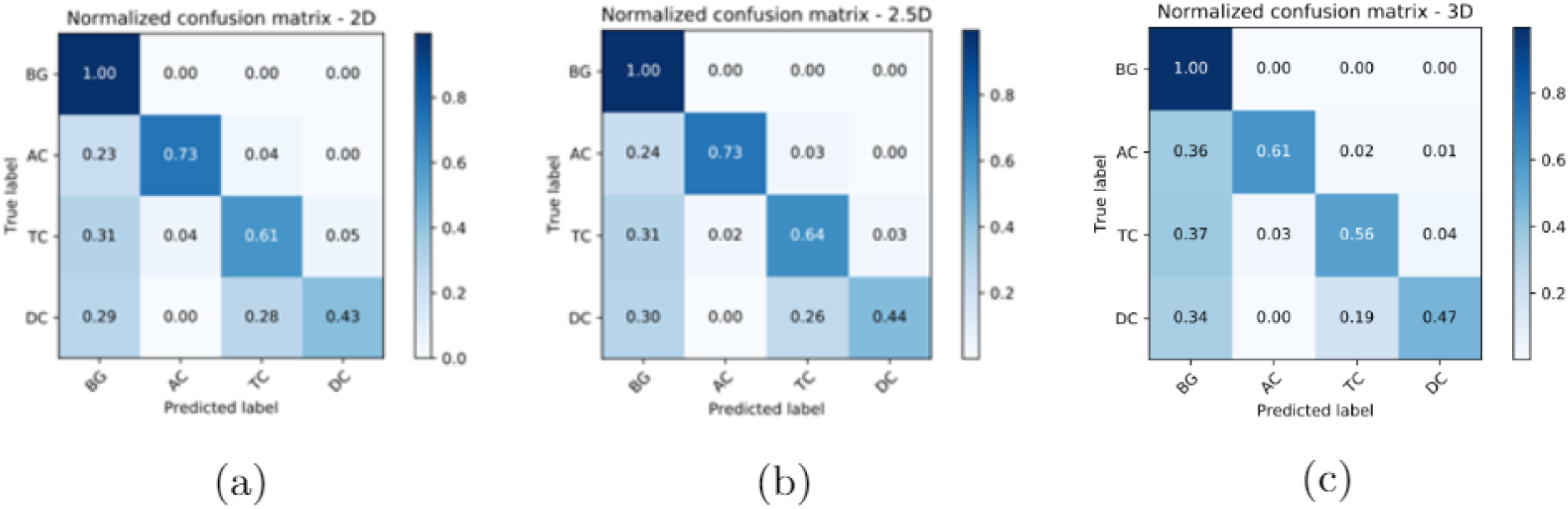
Class confusion matrices for 2D (a), 2.5D (b) and 3D (c) for the GE Medical Systems HDxt scanner. Each has been normalized and shows the percentage of class confusion for each predicted and true label.

**Figure 14:**
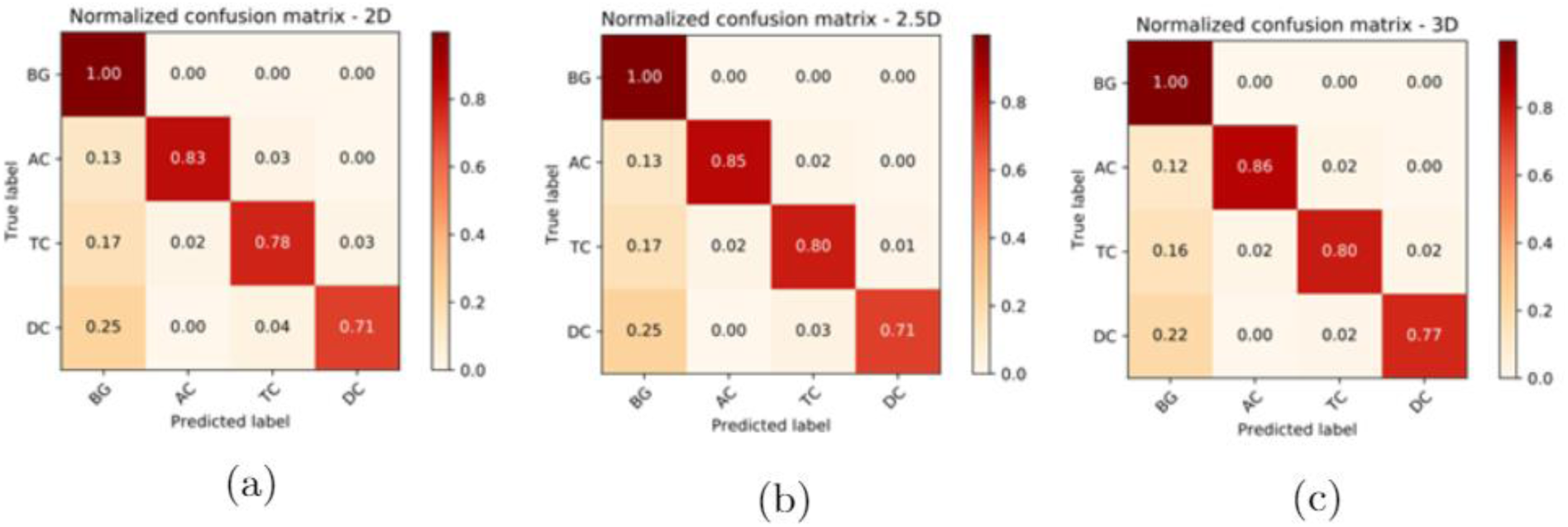
Class confusion matrices for 2D (a), 2.5D (b) and 3D (c) Philips Achieva scanner. Each has been normalized and shows the percentage of class confusion for each predicted and true label.

The class confusion pattern between colon sections remains the same between the two scanners. Misclassification between DC and TC is the highest error especially in the GE dataset.

## Sources of error

Figures 8, 9 and 10 show that the largest source of error is likely to be FG-BG class confusion caused by inaccurate boundaries, but there are instances of inter-class confusion which can account for individual class errors as can be see in Figure 9 with TC-DC class confusion. The confusion matrices in Figure 13 indicate that the most likely inter-colon class confusion is between the TC and DC sections. This is closely followed by misclassifying the TC as the AC and vice versa. These results are expected because the TC intersects both of the other two colon regions whereas the ascending and descending sections appear on opposite sides of the body. All networks have very few confusions between AC and DC. The remaining error comes from the misclassification of BG as FG and vice-versa. We find that the section which is most commonly confused with the BG is the DC (around 25% in each dataset).

### 2.5D and 3D Data Ablation

In order to assess the trained models’ performance and to gain insight into the inner workings of the network, we added ablations to the image volumes by ‘greying out’ regions using the mean intensity of the dataset. We hypothesised that these ablation experiments would show to what extent the models learnt to use 3D context information. The networks were not trained on these ablated sequences; instead, we use them as input at inference time. The aim was to examine how effective the networks were at filling in missing data based on surrounding context in the volume - hence it was not appropriate to test the 2D network this way. Data used in the experiments were taken from the test set to provide a fair evaluation. First, the middle slice in every triplet in the test set was ablated for the 2.5D network.

The 3D network received multiple instances of the same volume where only one slice is ablated, but the slice which was ablated was shifted a single place between instances. Overall the same slices which were ablated in the 2.5D network input were also ablated here. In a separate experiment, the 3D network received a single instance of each test set volume where every other slice was ablated. To ensure there was no bias we repeated this experiment with an offset of 0 and 1 so that all slices in a volume were ablated at least once. We refer to the two 3D experiments as single ablation and multi-ablation. The score for each volume in the test set was calculated as the average dice score on the ablated image in the volumes. In the multi-ablation experiments we averaged the results across the two offsets.

Table 4 shows the results of ablating images on the test set acquired from the Philips Achieva scanner. The 3D network outperformed the 2.5D network during single ablation by a significant Dice margin of 0.258 when carrying out binary classification. On the task of individual colon section segmentation, the 2.5D network is outperformed by 0.440. Overall, the accuracies obtained by the 2.5D network indicate that there was not a large amount of context information being used to segment the slices, and the 2.5D network appears to heavily rely on the main slice being segmented. The 3D network can use context from further afield in the volume, so potentially more context information is available to help fill in the ablated data.

**Table 4:**
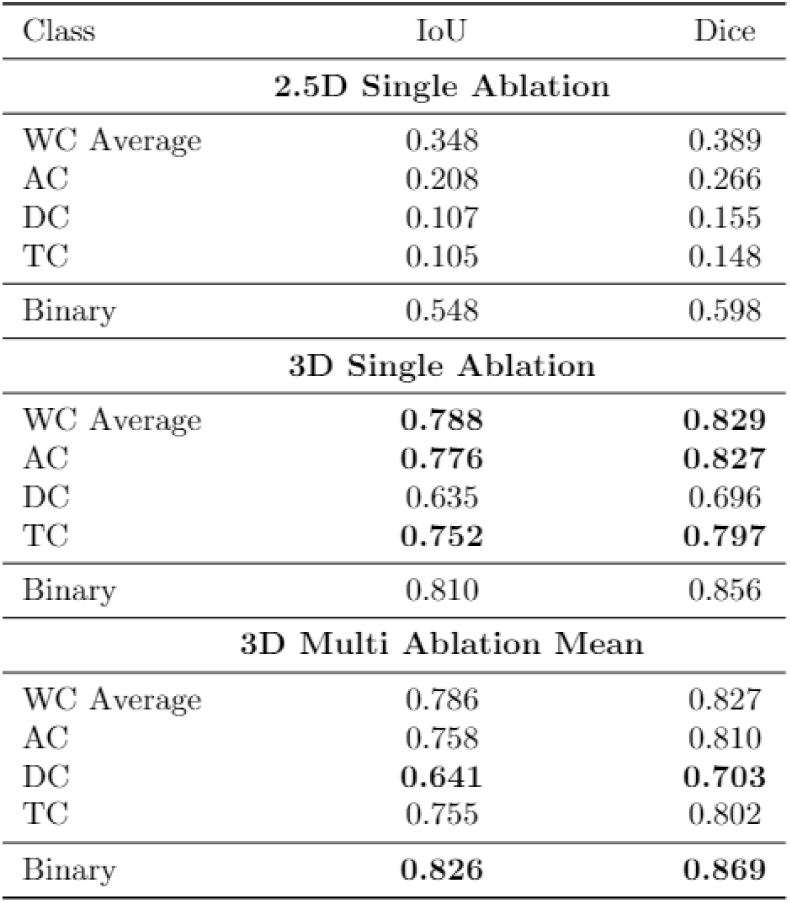
Results obtained for 2.5D and 3D networks on ablated test images from the Philips Achieva scanner

For the task of segmenting individual colon sections we found a minimal difference between the multi-ablation and single ablation experiments. Overall single ablation only yielded a Dice score 0.002 higher for WC Average labelling, however on the task of binary segmentation it achieved a score which was 0.013 lower than its multi-ablation counterpart. Single ablation achieved a higher accuracy for all colon sections except the descending colon. It is not clear why this occurred as we would expect the single ablation results to be higher for all sections of the colon; that said, the difference is only small and not considered significant. Comparing the performance of the network 3D network with and without ablation (Table 3 and 4), ablation only decreased the WC average Dice score of the 3D network by 0.069, whereas for the task of binary segmentation ablation decreases the dice score by 0.064

## Inference Time

Once models were fully trained their run times were compared, for the forward pass which is the equivalent to having the network give segmentation predictions. We found that the fastest network at inference time was the 2D network, followed by 2.5D and lastly 3D. Table 5 shows that the same trend was found when the data loading is included in the inference time. Distinguishing the effect of data load is important because typically for the 2D network only the current batch is loaded into memory. Since the number of slices in a volume is larger than the batch size to process the whole volume several loading sequences are required. This gives the 2D and 2.5D network a speed disadvantage.

**Table 5:** Time taken, in seconds, for each network to make predictions on a single volume from a Phillips achieva scanner

| Timing Method | 2D | 2.5D | 3D |
| --- | --- | --- | --- |
| Including Data Load(s) | 0.205 | 0.225 | 0.253 |
| Excluding Data Load(s) | 0.095 | 0.106 | 0.216 |

## CONCLUSIONS

In this paper, we have created effective CNNs for the automatic segmentation of the colon from MRI images of the abdomen. The 3D network performed better than the 2.5D network which in turn outperformed the 2D network for the task of segmenting individual colon sections. This indicates that depth information provides important context for a CNN. However on the task of binary segmentation context is less important, and we observes that the difference in performance between networks is much lower, with the 3D network performing best followed by 2D, and 2.5D networks.

However 2.5D networks may provide a better and computationally cheaper alternative if powerful hardware is not available and provides a faster network at inference time. 3D networks can be used in situations where powerful hardware is available and the increase in accuracy is deemed worth the cost.

We also showed that it is possible to use the 3D network successfully on images from an entirely different scanner without the use of transfer learning. The 2D and 2.5D networks struggled to segment volumes from other scanners.

Future work should seek to identify how well 3D networks perform with varying amounts of depth in a volume in order to identify the most optimal volume input size for a network for a particular task. Reducing volume size will alleviate the VRAM issues associated with volumetric convolutions. Future work should also focus on generalised networks able to deal with volumes from a range of scanners whilst minimising performance loss.

In conclusion, it has been shown that 3D CNNs are capable of accurately defining colonic volumes and may pave the way to the use of this MRI measurement in routine clinical practice for functional GI disorders, as well as expanding its use in the research setting.

## Data Availability

Data is not available at this time

